# Precision Digital Intervention for Depression Based on Social Rhythm Principles Adds Significantly to Outpatient Treatment

**DOI:** 10.1101/2022.05.06.22274565

**Authors:** Ellen Frank, Meredith Wallace, Mark J. Matthews, Jeremy Kendrick, Jeremy Leach, Tara Moore, Gabriel Aranovich, Tanzeem Choudhury, Nirav R. Shah, Zeenia Framroze, Greg Posey, Samuel Burgess, David J. Kupfer

**Affiliations:** Department of Psychiatry, University of Pittsburgh School of Medicine, Pittsburgh, PA, United States; HealthRhythms, Inc. Long Island City, NY, United States; School of Computer Science, University College, Dublin, Ireland; Huntsman Mental Health Institute. University of Utah School of Medicine, Salt Lake City, UT, United States; Cornell Tech, New York, NY, United States; Stanford University School of Medicine, Palo Alto, CA, United States; Sharecare, Atlanta, GA, United States

**Author notes:** **Correspondence:** Ellen Frank, PhD.

**Keywords:** depression, treatment, digital intervention platform, passive monitoring, depressive symtpoms, social rhythm disruption, social rhythm regularity

## Abstract

We conducted a 16-week randomized controlled trial in psychiatric outpatients with a lifetime diagnosis of a mood and/or anxiety disorder to measure the impact of a first of its kind precision digital intervention based on social rhythm regulation principles (Cue). The full intent to treat (ITT) sample consisted of 133 individuals, aged 18 to 65. The primary sample of interest were individuals with moderately severe to severe depression at study entry (baseline PHQ-8 score greater than or equal to 15; N=28).

Cue is a novel digital intervention platform that capitalizes on the smartphone’s ability to continuously monitor depression relevant behavior patterns and use each patient’s behavioral data to provide timely, personalized micro interventions, making this the first example of a digital precision intervention that we are aware of. Participants were randomly allocated to receive Cue plus care as usual or digital monitoring only plus care as usual.

Within the depressed and full ITT samples, we fit a mixed effects model to test for group differences in the slope of depressive symptoms over 16 weeks. To account for the nonlinear trajectory with more flexibility, we also fit a mixed effects model considering week as a categorical variable and used the resulting estimates to test the group difference in PHQ change from baseline to 16 weeks.

In the depressed at entry sample, we found evidence for benefit of Cue. The large group difference in the slope of PHQ-8 (Cohen’s *d*= -0.72) indicated a meaningfully more rapid rate of improvement in the intervention group than in the control group. The Cue group also demonstrated significantly greater improvement in PHQ-8 from baseline to 16 weeks (p=0.009). In the full sample, the group difference in the slope of PHQ-8 was negligible (Cohen’s *d*=-0.10); however, the Cue group again demonstrated significantly greater improvement from baseline to 16 weeks (p=0.040).

We are encouraged by the size of the intervention effect in those who were acutely ill at baseline and by the finding that across all participants, 80% of whom were receiving pharmacotherapy, we observed significant benefit of Cue at 16 weeks of treatment. These findings suggest that a social rhythm focused digital intervention platform may represent a useful and accessible adjunct to antidepressant treatment.

(https://clinicaltrials.gov/ct2/show/NCT03152864?term=ellen+frank&draw=2&rank=3)

## 1 Introduction

The chronic, global problem of inadequate access to mental healthcare has been described by many. However, the lack of accurate, consistent measurement in mental health has historically received less attention. Subjective, episodic collection of patient and clinician-reported survey data has been the only option available to clinicians since the field began. Digital behavioral phenotyping uses data collected from smart devices to create a personalized and precise picture of an individual’s behavior. For the first time, shortcomings of the legacy approach to measurement are avoided, by enabling continuous, objective, ecologically valid measurement (1). In addition to behavioral phenotyping, digital interventions, such as computerized versions of evidence-based approaches like cognitive therapy, have been developed to improve access to care and augment standard treatment (2).

We report on a randomized clinical trial of a digital phenotyping-based intervention platform for individuals with depression. To the best of our knowledge is the first example of a ‘precision digital intervention,’ i.e., it leverages the continuous data produced by the smartphone to generate inferences about the patient’s clinical state and surface the evidence-based micro-intervention that is most relevant to that patient at that moment – the right care at the right time.

Cue represents the first digital intervention based on the social rhythm regulation conceptual model (6,7). According to this model, chaotic, irregular daily routines are associated with the onset and maintenance of mental disorder symptomatology (8,9). A therapeutic approach that helps patients achieve more regular routines has been shown to improve mood disorder symptoms and maintain an asymptomatic state (10-15). We viewed the commercial smartphone as the ideal vehicle for measuring the extent of social rhythm disruption patients might be experiencing and for providing just-in-time micro-interventions to move them in the direction of greater social rhythm regularity.

Originally conceptualized as a feasibility and acceptability study, the National Institute of Mental Health encouraged the investigators to use the planned clinical population (psychiatric outpatients with a lifetime diagnosis of a depressive and/or anxiety disorder and any level of current symptomatology) to explore the potential efficacy of such a digital intervention platform. To do so, we compare an intervention platform that consisted of smartphone sensor-based digital monitoring, psychoeducation about the social rhythm regulation model and micro-interventions to aid patients in achieving increased social rhythm regularity (Cue) with sensor-based monitoring alone in a group of individuals who were concomitantly receiving outpatient psychiatric care as usual. We hypothesized that such a digital intervention platform would be associated with greater improvement in depressive symptoms than the comparator condition consisting of smartphone-based behavioral monitoring alone.

## 2 Materials and Methods

### 2.1 Study Designs and Participants

Outpatients being treated at the University of Utah Department of Psychiatry who had indicated at the outset of treatment that they were willing to be approached in the future about participating in research studies were contacted by phone on a sequential basis. Patients who appeared to be eligible and indicated an interest in the study were brought to the Department of Psychiatry for a single in-person visit to confirm eligibility and to obtain informed consent. All future contact with study participants occurred via email, text, or phone.

Patients were included if they had a lifetime diagnosis of a mood or anxiety disorder as assessed by the Mini International Neuropsychiatric Interview (16) were currently receiving treatment for their psychiatric disorder, were age of 16-65, if on psychotropic medication, were on a stable dose, owned and used an iPhone (5s or later) and were able and willing to provide written informed consent. Patients were excluded if they had a lifetime diagnosis of schizophrenia, antisocial personality disorder, primary obsessive-compulsive disorder, were currently psychotic or actively suicidal, met criteria for *current* alcohol, substance use or eating disorder. Those with a lifetime diagnosis of a bipolar disorder were not excluded if they were not currently psychotic. Individuals who had a poorly controlled medical condition that might cause confounding depressive symptoms (e.g., untreated thyroid disorder or lupus) or required medications that could cause depressive symptoms (e.g., high doses of beta blockers or alpha interferon were also excluded. Individuals who were taking medication at the time of study entry and required a change in dose or in medication(s) prescribed were discontinued from the protocol at the time of medication change. All participants provided informed consent according to procedures approved by the University of Utah Institutional Review Board.

Participants downloaded either the monitoring only app which was used to collect both passively sensed data (pedometer, display status, GPS location) and self-reported measures of clinical status or the full digital intervention platform consisting of: 1) the monitoring component, 2) a series of psychoeducational Learning Modules and 3) personalized micro-interventions that were selected based on the user’s sensed behavioral data. Following download, participants were contacted by study personnel (typically only via email) only if there appeared to be a problem in the receipt of their sensed or self-reported data.

### 2.2 Randomization and Patient Allocation

Participants were allocated on a 1:1 basis to either the control condition that consisted of treatment as usual by University of Utah faculty psychiatrists plus passive monitoring only or to an experimental group that received treatment as usual plus monitoring, psychoeducation, and personalized micro-interventions for a period of 16 weeks. The micro-interventions were pushed to patients through their phones on a two to three times per week basis (see Figure 1).

**Figure 1.**
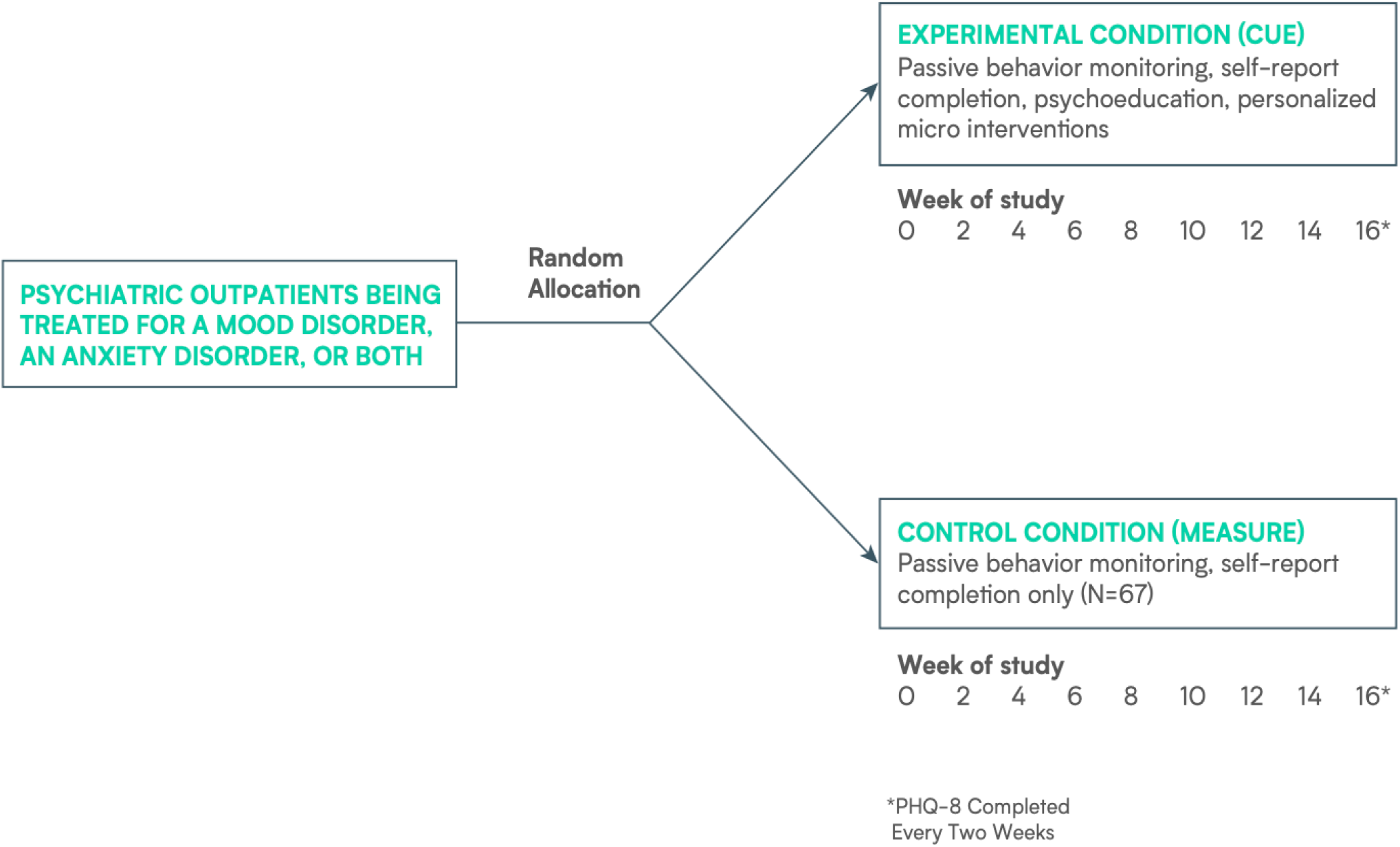
Design of RCT Comparing a Digital Intervention Platform with Monitoring Only.

### 2.3 Experimental Intervention

The Cue digital intervention platform continuously monitors behavioral biomarkers and collects patient-reported data that provide an additional measurement of patients’ daily routines or ‘social rhythms’ such as sleep timing and daily self-rating of the user’s mood and energy.

At the outset of the treatment period, the Cue platform provides patients with psychoeducational material, referred to as Learning Screens. The Learning Screens provide the rationale for the regular routines that the intervention is intended to help patients achieve and why such regularity can help reduce the severity of depressive symptoms and maintain mood stability. The 10 Learning Modules are released to the patient on a timed basis over the first three weeks of treatment but are also available for patients to refer to later if the patient chooses to review them (see Figure 2).

**Figure 2.**
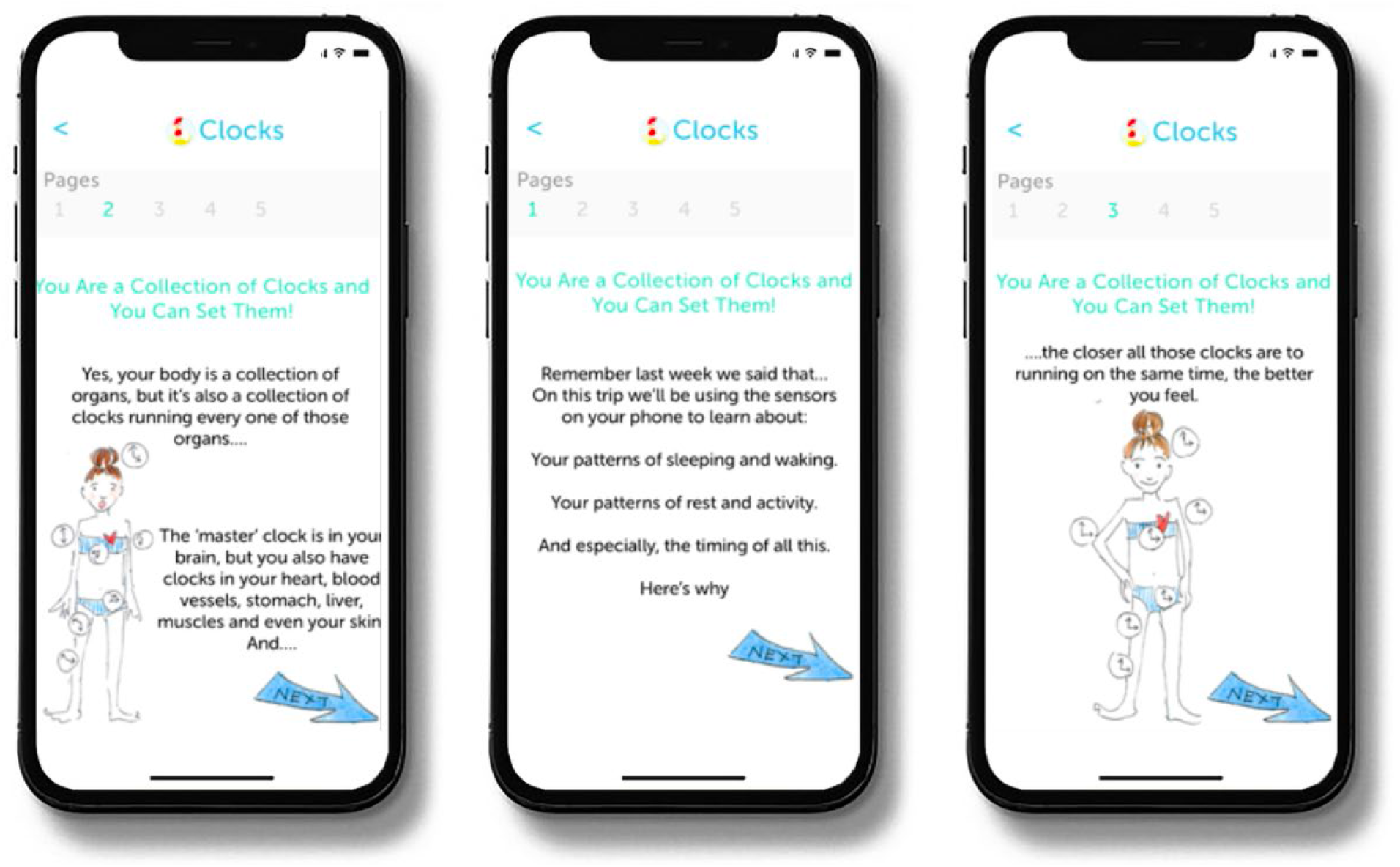
Sample Learning Screens.

The digital intervention platform evaluated here is built on a well-validated conceptual framework (6,7) that links irregularity of behavioral rhythms to illness onset (8,9) and stability of behavioral rhythms to recovery and sustained wellness (10-15). Using this conceptual framework, the platform interprets data gathered through phone sensors and patient self-reports to generate micro-interventions to improve patients’ depression symptoms. In the present study, the intervention platform followed a 16-week timeline for providing such micro-interventions that were sent as push notifications to the patient’s phone approximately every two to three days. An example of such a behavior change suggestion is displayed in Figure 3.

**Figure 3.**
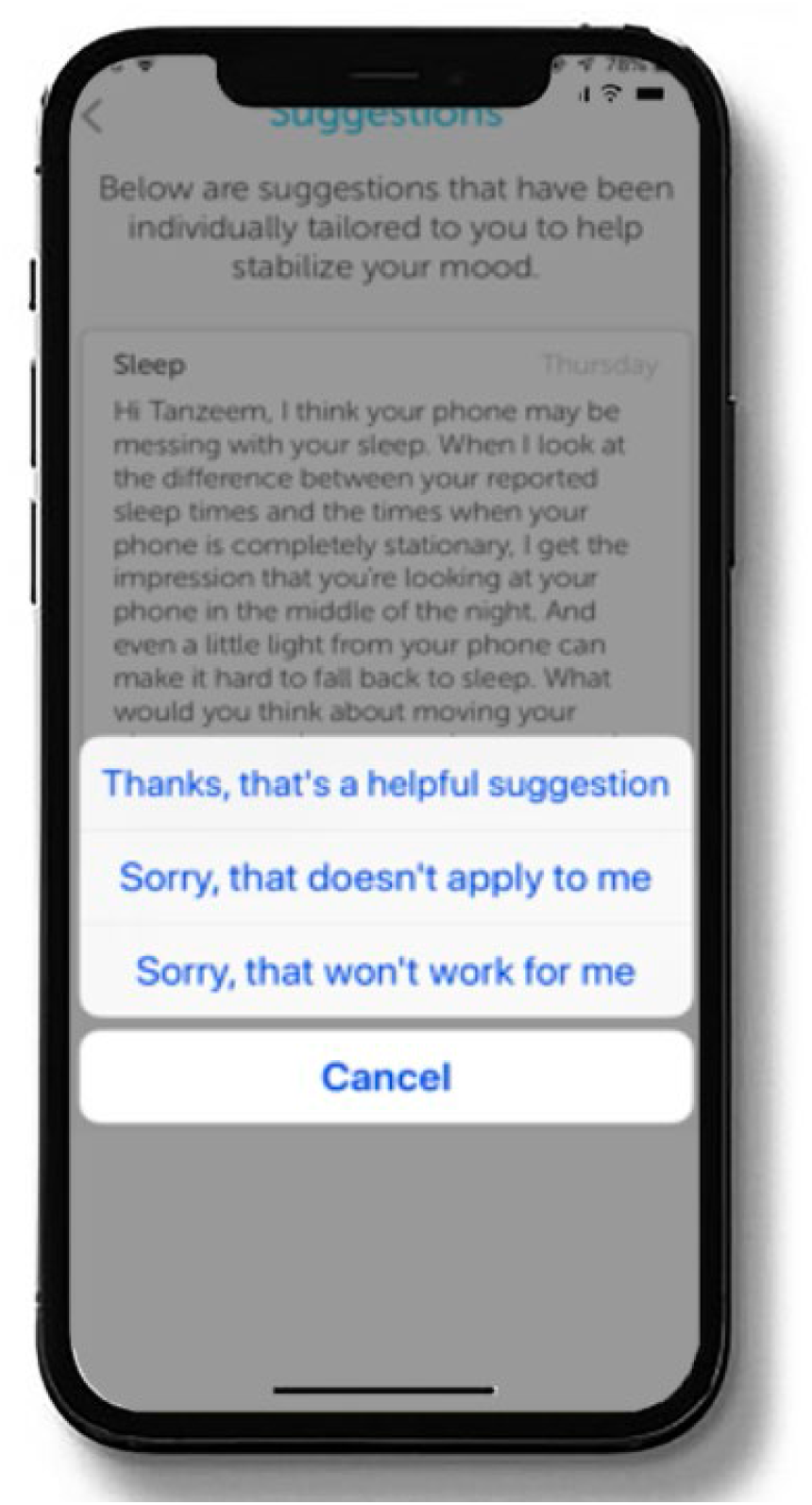
Sample Micro-Intervention/Behavior Change Suggestion.

All participants completed a daily rating of mood on a visual analogue scale (scored from 0=worst I’ve ever felt to 10=best I’ve ever felt) and the PHQ-8, a well-accepted measure of depression severity (17) every two weeks throughout the study. We chose to use the PHQ-8 rather than the PHQ-9 which includes a suicidality item because the entirely virtual nature of the study did not allow for adequately rapid attention to reports of suicidality. Study participants were compensated on an escalating basis with gift cards valued at $50, $75, $100, and $125 for the completion of the self-report questionnaires after each month of participation.

In the present study, each participant’s sensed behavioral and self-reported data were uploaded continuously to the HealthRhythms servers and then summarized on a dashboard that enabled an ongoing record of patient status. Personalized micro-interventions were crafted by the first author and Ms. Moore, both of whom are well-versed in the social rhythm regulation model. Reading of these personalized micro-interventions required minimal patient time (generally between 30 seconds and one minute) and reflected what was currently happening in the individual patient’s life as indicated in their sensed data. For example, a patient who has a regular wake time during the week but tends to ‘sleep in’ on Saturdays and Sundays might receive a micro-intervention pointing this out, reminding the patient that one of the best ways to keep the body’s clock running efficiently is to stick to approximately the same wake time seven days a week. To facilitate this behavior change, the micro-intervention would typically suggest a means of achieving this goal such as considering meeting a friend for a walk or a coffee at 8:00AM on Saturday and Sunday, so that their weekend wake times would approximate their weekday wake times. The micro-intervention would then encourage the patient to do so by pointing out that they will probably find it easier to get up on time and feel better on Monday if they try this.

### 2.4 Control Intervention

The control condition consisted of Measure, a digital platform for sensed and self-reported behavioral monitoring. Participants allocated to the Measure monitoring only condition completed the same daily and bi-weekly self-report measures as those allocated to the experimental condition but did not receive any of the psychoeducational material or any behavior change micro-interventions.

### 2.5 Full Study Sample

We consented 135 patients with a lifetime diagnosis of a mood and/or anxiety disorder to the 16-week trial. Two participants did not receive the allocated intervention, leaving 133 analyzable participants for the full study. Ten patients (7.4%) dropped out of the study prior to study week 16, and 4 (3%) were discontinued from the study according to the protocol because they experienced a change in medication (see Figure 4). The 133 analyzed participants were between the ages of 18 and 65, with a mean age of 33(±11). Seventy-four percent (n=98) were female. Nineteen percent (n=25) had a lifetime mood disorder diagnosis only, 11% (n=15) had a lifetime anxiety diagnosis only, and 70% (n=93) met lifetime criteria for both a mood disorder and an anxiety disorder. Eighty percent (n=107) of participants were receiving pharmacotherapy prescribed by a University of Utah Department of Psychiatry physician (see Table 1). Data quality was excellent; the median coverage of 24/7 sensed behavioral data was 99.09% across all study days and the mean was 93.32%.

**Figure 4.**
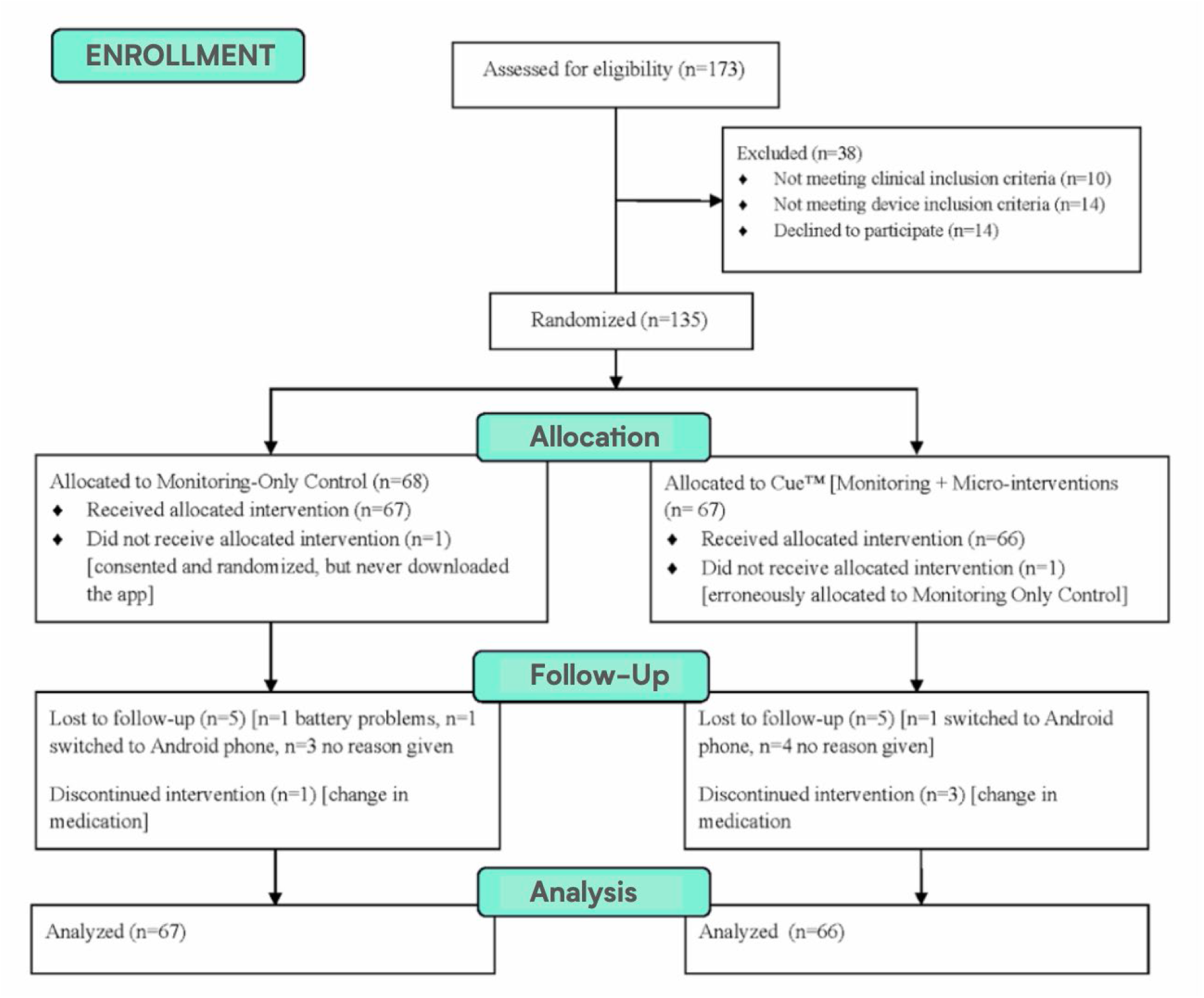
Consort Diagram.

**Table 1.** Demographic and Clinical Characteristics for the Full ITT Sample - Cells show Mean (SD) for continuous and % (N) for categorical measures

|  | All (N=133) | Cue (N=66) | Measure (N=67) | Comparison |
| --- | --- | --- | --- | --- |
| Sociodemographic Factors |  |  |  |  |
| Age | 32.94(11.39) | 33.85(11.64) | 32.04(11.15) | t = 0.91, p = 0.36 |
| Female | 73.68(98) | 72.73(48) | 74.63(50) | $\chi^2$< 0.01, p = 0.96 |
| Employment Status | | | | $\chi^2$= 2.38, p = 0.31 |
| Full Time Work | 60.15(80) | 65.15(43) | 55.22(37) |  |
| Part Time Work | 25.56(34) | 19.7(13) | 31.34(21) |  |
| Not Working | 14.29 (19) | 15.15(10) | 13.43(9) |  |
| Living Situation | | | | $\chi^2$= 5.38, p = 0.07 |
| Living Alone | 17.29(23) | 22.73(15) | 11.94(8) |  |
| Living with Family | 63.91(85) | 65.15(43) | 62.69(42) |  |
| Living with Unrelated People | 18.8(25) | 12.12(8) | 25.37(17) |  |
| Clinical Characteristics |  |  |  |  |
| First PHQ | 11.02(4.74) | 11.53(5.14) | 10.52(4.28) | t = 1.23, p = 0.22 |
| First PHQ ≥15 | 21.05(28) | 24.24(16) | 17.91(12) | $\chi^2$= 0.47, p = 0.49 |
| Depression or anxiety medication | 80.45(107) | 80.3(53) | 80.6(54) | $\chi^2$< 0.01, p > 0.99 |
| Lifetime diagnosis | | | | $\chi^2$= 1.97, p = 0.37 |
| Mood disorder only | 18.8(25) | 18.18(12) | 19.4(13) |  |
| Anxiety disorder only | 11.28(15) | 7.58(5) | 14.93(10) |  |
| Mood and Anxiety disorder | 69.92(93) | 74.24(49) | 65.67(44) |  |

### 2.6 Patients Entering in a Major Depressive Episode

To assess the effect of the digital intervention on individuals suffering from a fully symptomatic episode of major depression at study entry, we selected those individuals who scored >=15 on the PHQ-8 at their baseline assessment, an indicator of moderately severe or severe depression (17). This group consisted of 28 individuals, 12 of whom were allocated to monitoring only and 16 of whom were allocated to the full digital intervention platform. Twenty-five (89%) of these individuals were receiving antidepressant pharmacotherapy prescribed by University of Utah faculty psychiatrists at the time of study entry. Twenty-seven of these 28 participants completed the full 16-week trial. The one non-completer was withdrawn per protocol because of a change in medication, leaving 27 analyzable participants (see Table 2).

**Table 2.**
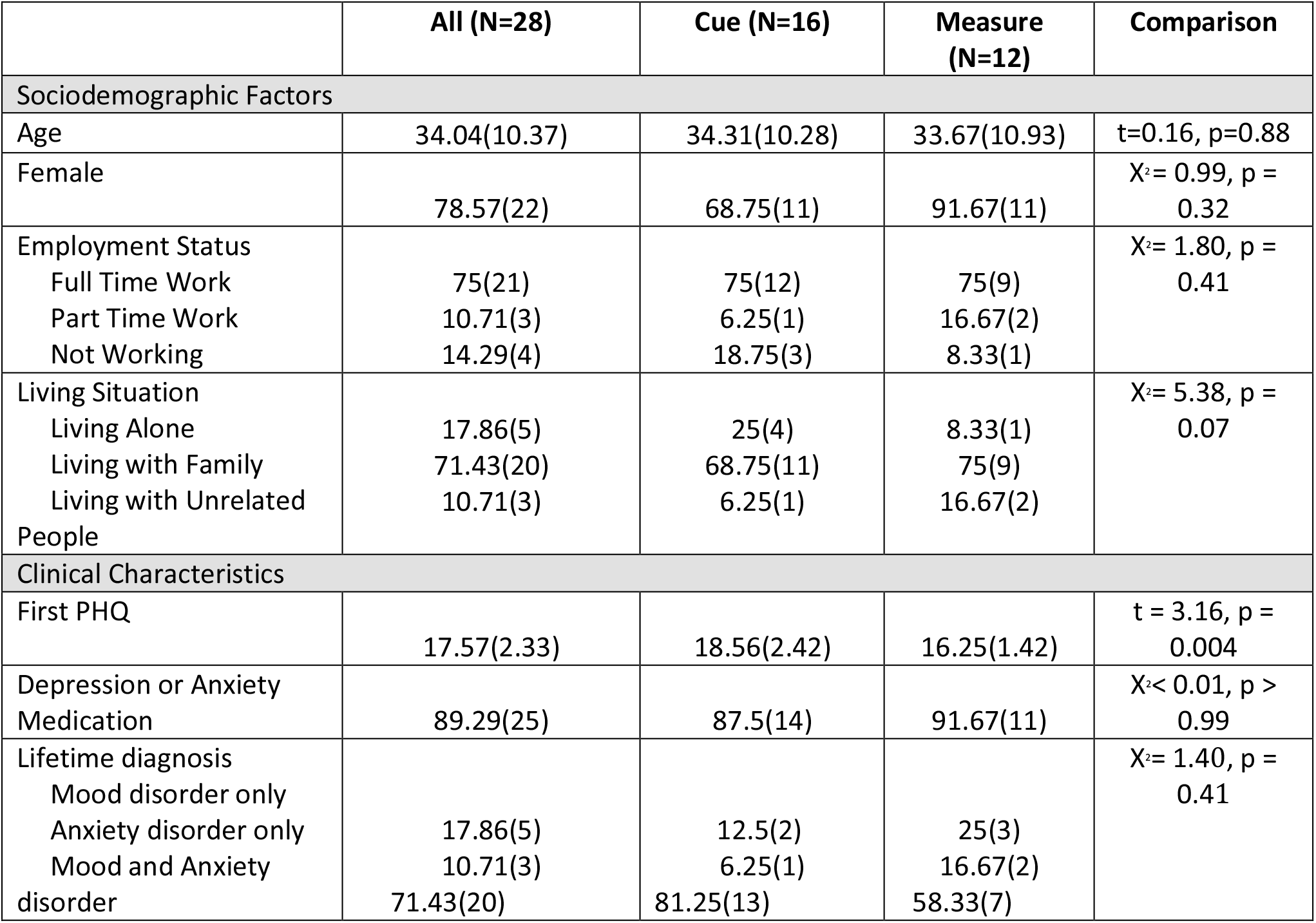
Demographic and Clinical Characteristics for the Depressed-at-Entry ITT Subsample (Baseline PHQ≥15) Cells show Mean (SD) for continuous and % (N) for categorical measures.

### 2.7 Outcome analysis

In the depressed-at-entry sample (PHQ-8 ≥15; N=28) and then in the full study sample (N=133), we fit a mixed effects model with a random intercept and slope to test for group (experimental vs. control) differences in the slope of depressive symptoms over 16 weeks. Because there was a non-linear effect of week such that initial reductions in PHQ-8 were steeper than those later in follow-up, we used a square-root transformation of week. Evaluation of model fit criteria (AIC, thBIC) indicated this was an optimal transformation (as compared to no transformation or a log transformation). Thus, these models included repeatedly measured PHQ-8 as the outcome with square root of week, group (Cue versus Measure), the square root week by group interaction, and covariates. Random intercept and time effects were included. From this model we also extracted the predicted random slope for each individual and used this to compute Cohen’s d effect sizes for group differences in slope over 16 weeks. Secondarily, to allow further flexibility in the trajectory of PHQ-8 over follow-up, we fit a mixed effects model with categorical time (baseline, 2, 4, 6, 8, 10, 12, 14, 16 weeks), group, and the categorical time by group interaction, with specific contrasts to test group differences in changes from baseline to 16 weeks. All analyses were conducted according to the principle of intention to treat (ITT). All models covaried for age, sex, living status, employment status, pharmacotherapy use, and lifetime diagnosis.

## 3 RESULTS

### 3.1 Depressed-at-Entry Sample

We focus first on the 28 study participants who entered the trial in a fully symptomatic episode of major depression, as these results have the greatest relevance for the ultimate utility of the Cue platform. In this subsample, the group by time interaction approached conventional levels of statistical significance (B[SE]=-1.18[0.62], p=0.059). Notably, however, the effect size of the group difference (Cue versus Measure) in slope was moderate to large (Cohen’s d [95% CI] =-0.72 [-1.53, 0.09]), indicating that individuals allocated to the experimental condition had a meaningfully steeper rate of improvement in PHQ-8 than those allocated to monitoring alone (see Figure 5). When treating time as categorical in secondary analyses, post-hoc analyses indicated a significant group difference in the PHQ-8 change from baseline to 16 weeks (Estimate [SE] = -4.74 [1.81], t=-2.63, p=0.009). See Figure 6.

**Figure 5.**
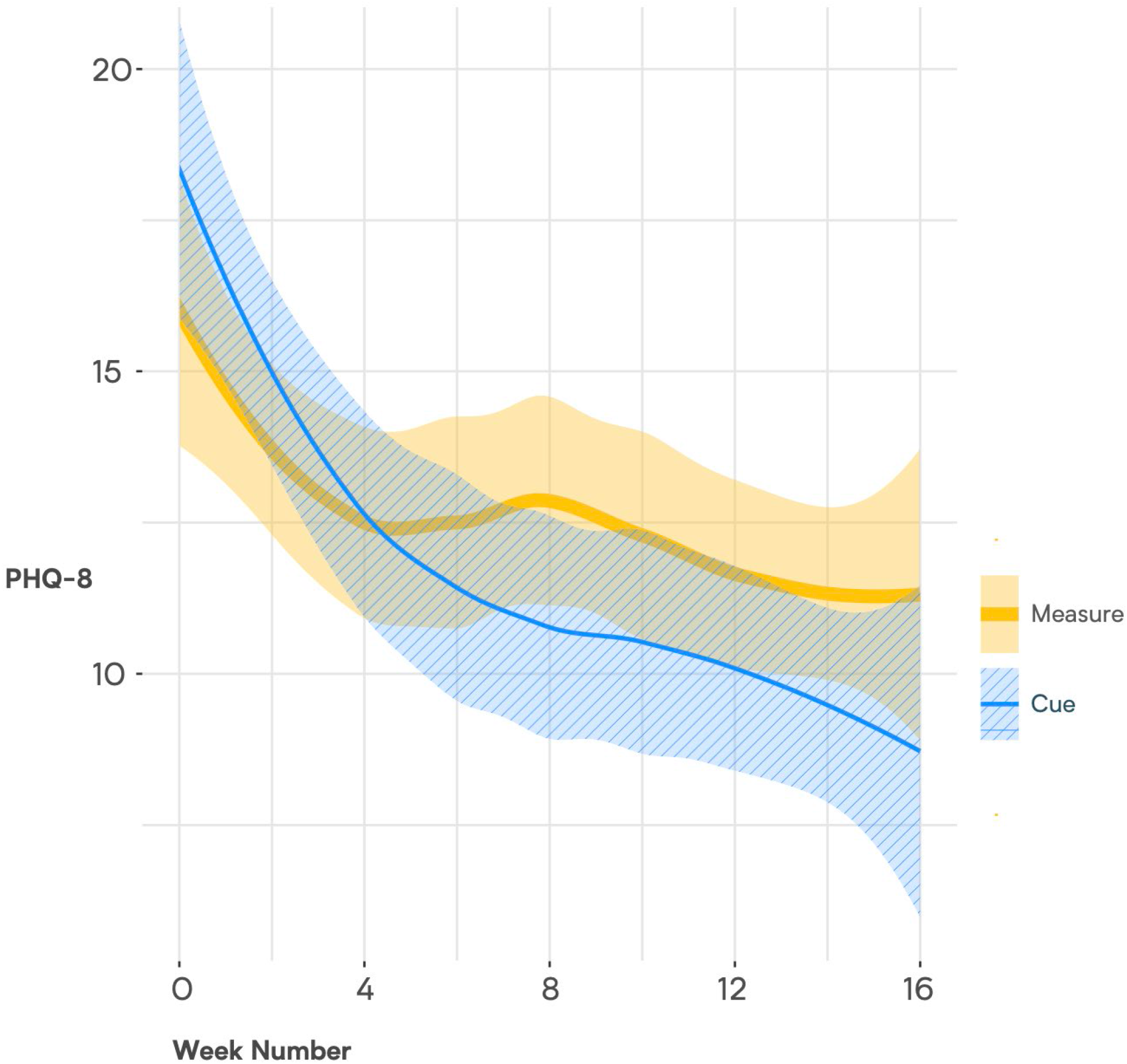
Loess Trajectories of PHQ-8 for Cue and Measure (control) Conditions in Depressed-at-Entry Participants (initial PHQ-8>= 15) Shaded areas represent the standard error around the estimated line.

**Figure 6.**
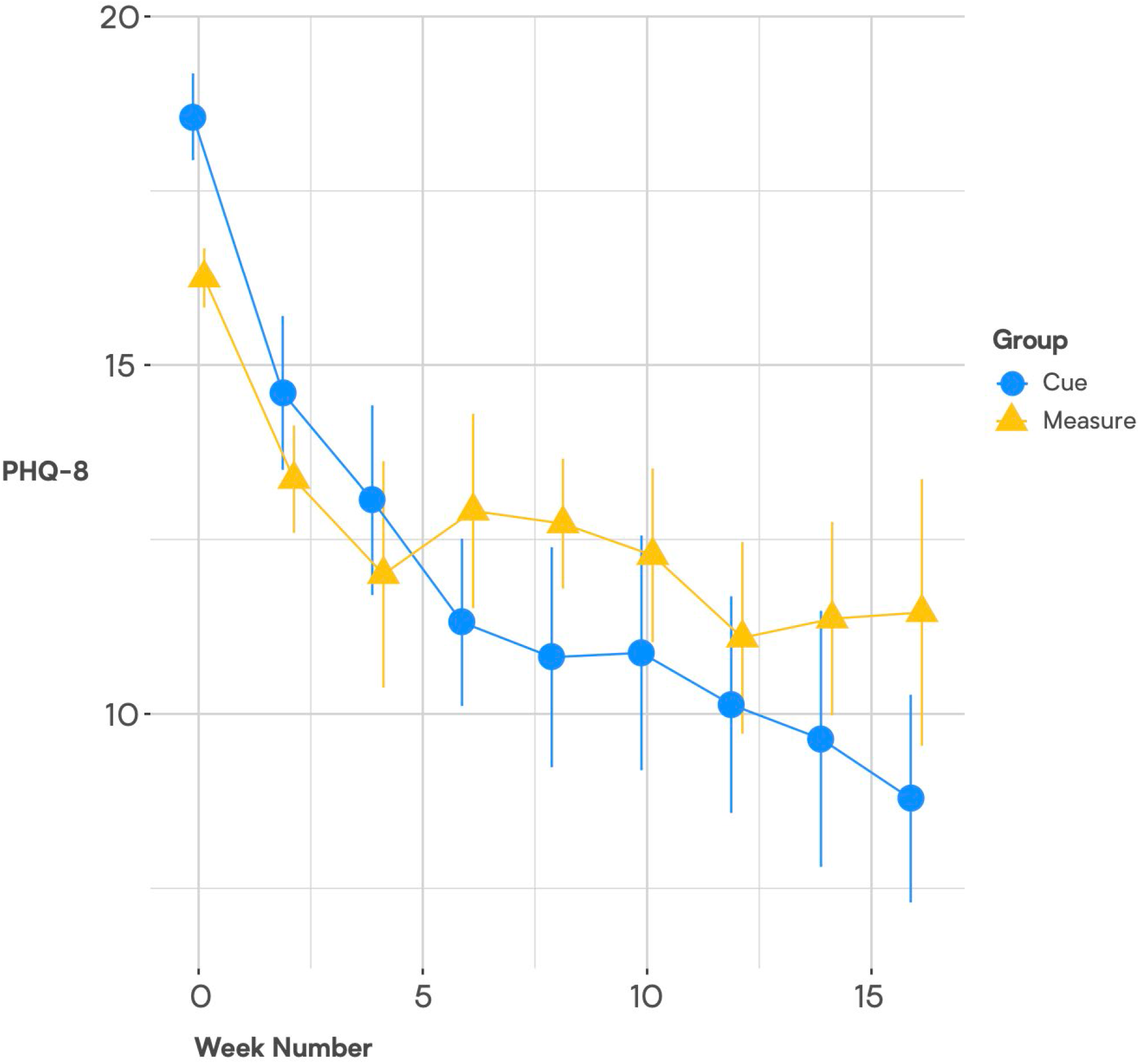
Means and Standard Errors of PHQ Scores by Study Week-Depressed-Entry-Sample.

### 3.2 Full Study Sample

In the full study sample, as indicated in Figure 7, the slopes of change in PHQ scores did not differ significantly for Cue versus Measure (B[SE]= -0.15 [0.22], p=0.507; Cohen’s d [95% CI] = -0.10 [-0.45, 0.24]. However, there were significant group differences in change from baseline to 16 weeks (Estimate [SE] = -1.50[0.73], p=0.040), with the group assigned to the digital intervention platform experiencing the greater improvement (See Figure 8)

**Figure 7.**
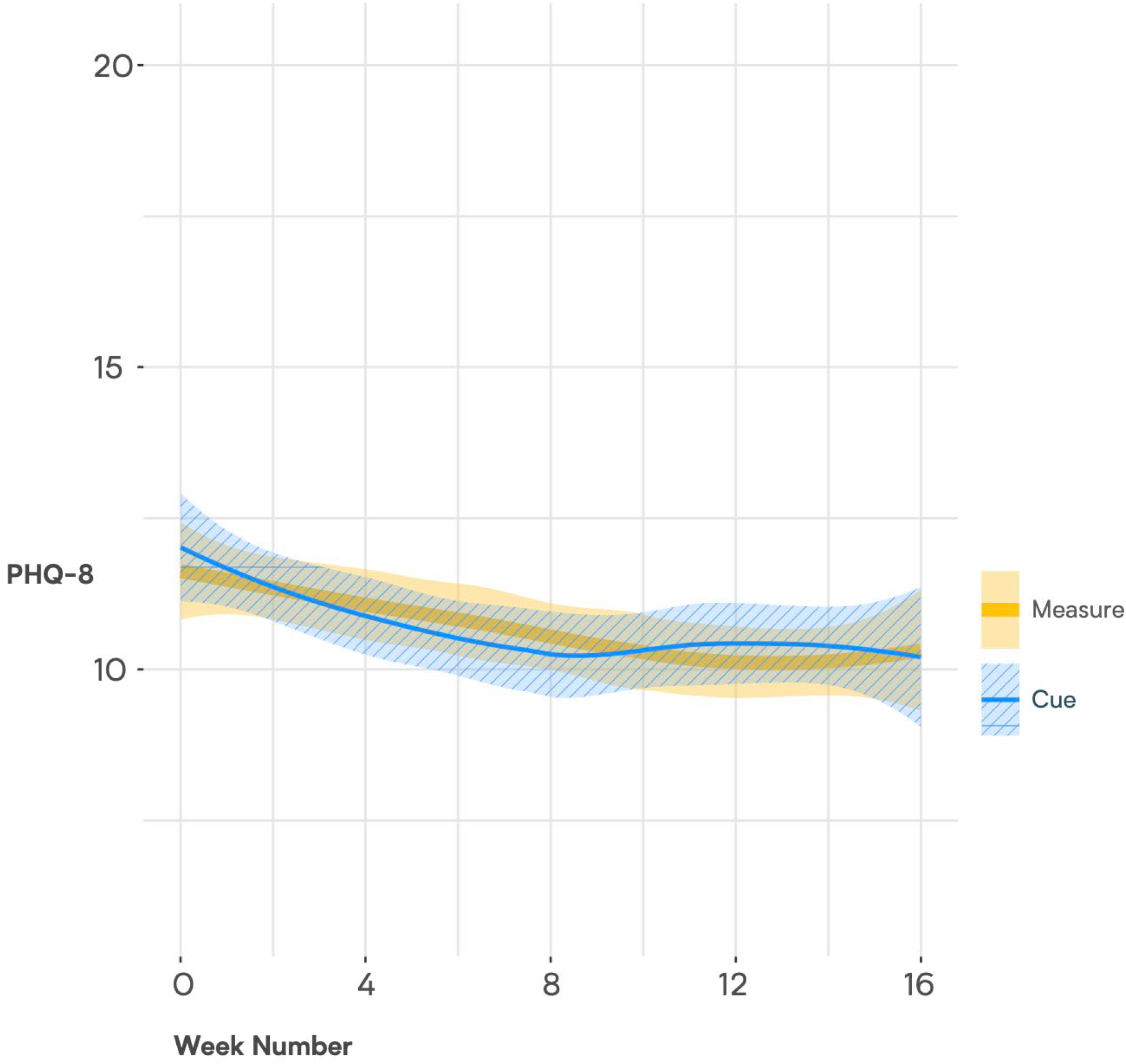
Loess Trajectories of PHQ-8 for Cue and Measure (control) Conditions in Full Study Sample. Shaded areas represent the standard error around the estimated line.

**Figure 8.**
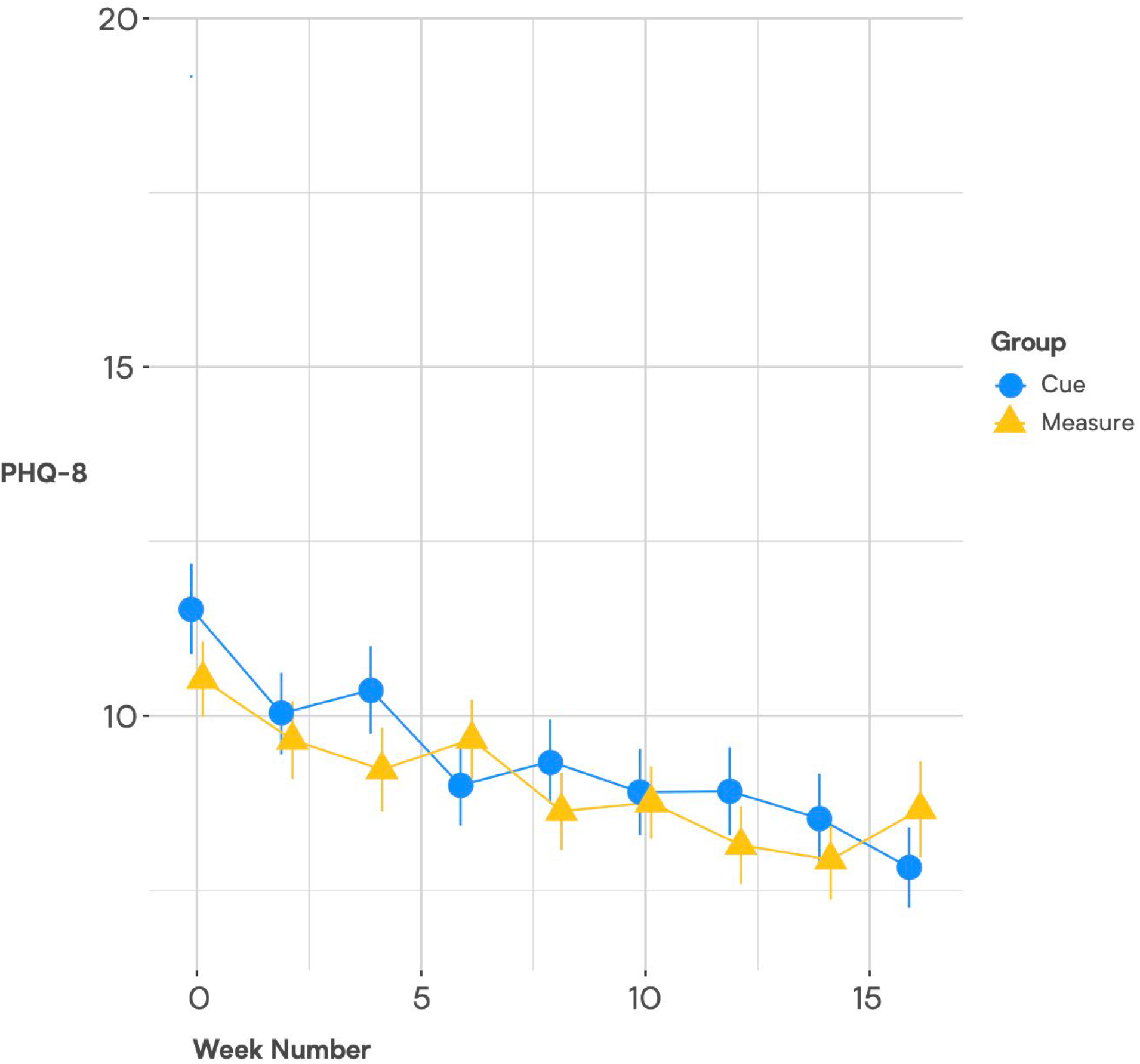
Means and Standard Errors of PHQ Scores by Study Week-Full Sample.

## 4 DISCUSSION

Among patients who were experiencing a moderately severe to severe episode of depression at baseline, we found evidence for significant benefit of a digital therapeutic based on social rhythm regulation principles when combined with outpatient psychiatric care consisting primarily of antidepressant pharmacotherapy. As part of a meta-analysis by Linardon et al. (18) reported on four trials that examined whether adding a smartphone intervention to a standard intervention (face-to-face, computerized, pharmacotherapy) for depression was superior to a standard intervention-only condition. The pooled effect size for the four comparisons between smartphone intervention + standard intervention vs. the standard intervention-only arm was g=0.26 (95% CI: –0.09 to 0.61). A recent meta-review of meta-analyses of digital interventions (19) reported that smartphone interventions for depression did not differ significantly…when tested as an adjunct to treatment (d = 0.26 [21]). The findings in the present report suggest that the effect size observed for Cue, when added to standard outpatient care in an academic department of psychiatry for patients with major depression, is substantially larger than those reported to date.

It is noteworthy that the pattern of change displayed in Figure 6 is typical of that observed in many studies of depression treatment, i.e., one in which there is initial improvement in both conditions probably as a result of the optimism and clinical attention associated with entering a treatment research program, which is then followed by a clinically meaningful separation between the active and control conditions.

In the full study sample, consisting of psychiatric outpatients with any level of symptomatology at baseline, we did not observe a significant difference in the slopes of change of the two groups. However, we did observe significant differences between the two study conditions in change in PHQ-8 scores from baseline to 16 weeks.

Several aspects of this trial are worthy of comment. It is noteworthy that the majority (80%) of study participants were receiving pharmacotherapy prescribed by University of Utah faculty psychiatrists throughout the course of the study. Thus, the observed effects of the digital intervention platform evaluated here are over and above the effects of standard of care pharmacotherapy. It has been very rare for any psychosocial intervention to add significantly to evidence-based pharmacotherapy. Indeed, the only instance of this of which we are aware was the study of Cognitive Behavioral-Analysis System of Psychotherapy (CBASP) in patients with chronic depression (20). This is particularly relevant because the context in which we anticipate Cue being used is likely to be one in which most users are receiving medication. It is also worth noting that proportion dropping out of the trial (7.4%) is among the lowest ever reported in a clinical trial of a depression intervention where drop-out rates in the 17.5% to 47.8% range are common (21), both Cooper & Conklin (21) and Torous et al (22) reported that dropout rates approaching 50% were typical of digital intervention trials for depression.

Much work has been done on the question of how best to deliver mHealth interventions. (23,24,25) Much has also been written on developing behavioral biomarkers for various conditions based on specific sensors and variables, such as sleep, social media engagement etc. (26-36) In contrast, we are not aware of any previous effort to combine the benefits continuous capture of objective behavioral biomarkers with a platform for the delivery of a digital intervention based on each user’s own behavioral data. Nor are we aware of another digital therapeutic based on social rhythm regulation principles, an approach to which smartphone monitoring is especially well-suited because these devices can capture rhythm-relevant behaviors in a completely passive manner on a continuous 24/7 basis. We believe this combination of the novel capacities of the commercial smartphone and a therapeutic model uniquely suited to those capacities may partially explain the high levels of treatment adherence and the positive outcomes observed.

The present study is limited in several respects. The study population consisting of psychiatric outpatients with any level of symptom severity at baseline was not ideal for addressing the question of the efficacy of the Cue platform; nonetheless, the subset of patients experiencing at least moderately severe depression at baseline provided an opportunity to explore the potential therapeutic effects of this approach to digital intervention. The relatively small number of participants in the depressed-at-entry sample represents another limitation; however, the large effect size observed in this subgroup is encouraging. Finally, the results reported here emerge from a clinical trial setting in which participants were compensated modestly for completion of the self-report instruments included and may not be generalizable to clinical practice. The amount of human attention given to the participants following the initial evaluation visit was minimal. Participants were only contacted by study personnel (typically only via email) if there appeared to be a problem in the receipt of their sensed or self-reported data and most participants had no contact with study personnel after the baseline/consent visit.

Encouraged by these results and recognizing that a platform that requires human curation of behavior change suggestions is not sustainable or scalable, we have developed an automated micro-intervention generation engine based on AI and ML principles that is currently being tested in an adaptation of Cue that is focused on patients with sleep problems. To support this effort, we have developed a library of over 1,500 behavior change suggestions to address various aspects of insomnia. Based on the individual patient’s sensed data, the platform uses AI to select and then send the appropriate micro-intervention for that user at that point in time. We will shortly be applying this automated approach to Cue for depression and are hopeful that these fully automated platforms will represent practical, scalable, and cost-effective approaches to addressing a variety of mental health problems.

## Data Availability

All data contained in the present report are available upon reasonable request to the authors

## 5 Contribution to the Field

Digital interventions represent one of the most promising approaches to increasing access to mental healthcare. Several examples of digital interventions have been developed and disseminated to patients via their computers and smartphones. These interventions benefit from their limitless, low-cost scalability, and evidence continues to grow that they are effective. However, they have historically been one-size-fits-all applications, which may account for a problematic record of poor engagement, with patients rapidly losing interest in static content.

Cue, a digital monitoring, psychoeducation, and intervention platform for depression, is the first example of a precision digital intervention. Cue’s AI-based solution translates passively acquired data from normal smartphone sensors into detailed, continuous behavioral information about the patient’s mental health status. Cue then uses that information to provide just-in-time micro-interventions sent to patients through the phone based on their own behavioral data, exposing patients to the intervention that is most relevant to them at any given moment.

We compared outcomes for Cue compared to behavioral monitoring alone among patients receiving treatment, primarily antidepressant medication, at the University of Utah Department of Psychiatry. We found that depressed patients who received Cue, in addition to outpatient treatment, improved approximately two times more than patients who received only outpatient treatment plus behavioral monitoring.

## 7 Author Contributions

EF, MM, TM, JK, TC and DK contributed to conception, design and conduct of the study. MM, GP, and SB developed the digital platforms to be tested. GP and JL organized the database. MW and JL performed the statistical analysis. EF, NS and ZF wrote the first draft of the manuscript. EF, MW, and GA wrote additional sections of the manuscript. All authors contributed to manuscript revision, read, and approved the submitted version.

## Acknowledgments

All authors have seen and approved the manuscript. The authors would like to acknowledge Allison Bustamante, Sarah Commesso, and Paul Gilbert of HealthRhythms, Inc. for their dedication and support of this work. They would also like to thank Adam Haim, PhD of the National Institute of Health for his support and encouragement throughout the process of developing and testing of the digital intervention described here.

## Supplementary Material

